# Machine learning in medicine using JavaScript: building web apps using TensorFlow.js for interpreting biomedical datasets

**DOI:** 10.1101/2023.06.21.23291717

**Authors:** Jorge Guerra Pires

## Abstract

Contributions to medicine may come from different areas, and most of these areas are filled with researchers eager to contribute. In this paper, we aim to contribute through the intersection of machine learning and web development. We employed TensorFlow.js, a JavaScript-based library, to model biomedical datasets using neural networks obtained from Kaggle. The principal aim of this study is to present the capabilities of TensorFlow.js and promote its utility in the development of sophisticated machine learning models customized for web-based applications. We modeled three datasets: diabetes detection, surgery complications, and heart failure. While Python and R currently dominate, JavaScript and its derivatives are rapidly gaining ground, offering comparable performance and additional features associated with JavaScript. Kaggle, the public platform from which we downloaded our datasets, provides an extensive collection of biomedical datasets. Therefore, readers can easily test our discussed methods by using the provided codes with minor adjustments on any case of their interest. The results demonstrate an accuracy of 92% for diabetes detection, almost 100% for surgery complications, and 80% for heart failure. The possibilities are vast, and we believe that this is an excellent option for researchers focusing on web applications, particularly in the field of medicine.

## 1. Introduction

Contributions to medicine may come from different areas; and most areas are full of researchers wanting to support. Physicists may help with theory, such as for nuclear medicine. Engineers with machineries, such as dialysis machine. Mathematicians with models, such as pharmacokinetics. And computer scientists with codes such as bioinformatics. We hope to contribute with machine learning for medical problems focused on web applications. More precisely, our intention is to present an accessible tool for constructing machine learning models, suitable for use by individuals without specialized expertise in the field.

Recently, thanks to several libraries, JavaScript became a viable option for designing machine learning. Nowadays, the most well-accepted and showing promising results is TensorFlow.js [1, 2, 3]. Thus, add all the advantages of working with JavaScript and machine learning in one web application. For our case, the core advantages are: i) the data will never leave the browser, ideal for sensitive data, most likely the case for medical information; ii) the calculations are done on the browser, no need to buy expensive server for scientific computations, ideal for startups and similar experimental endeavours. We believe that startups may support on decreasing the ever-growing costs of medical assistance [4]. They have already done that with text processing using artificial intelligence (e.g., chatGPT).

We have also added another ingredient to this solution: Angular [5]. Angular is a Single Page Application (SPA) creator, and we have already tested it for scientific computation [6]. For our case, the core advantage, on a possible real-scenario: since it is a SPA, the information will never leave the browser. When you have a server (e.g., Python), the user data will be sent to the server, and this is well-known to open space for attacks on sensitive data. In some medical protocols, you can not even store the patient data on a permanent driver, it is considered unethical [7, 8]. Talking to servers is also well-known to create delays. We had an experience in [6] with FASTA files that due to their big size, it was hard to keep sending those data around since HTTP calls have limits; we needed programming tricks to actually show the dataset at Machine learning in medicine using JavaScript the frontend.

Our goal here is showing the reader how easy it is to build powerful machine learning models using TensorFlow.js. Even though we know it is not straightforward to build models in medicine, we can arrive to promising results, with free libraries, and in seconds; of course, using model in real-scenario may require more care, such as making sure the dataset used is not biased against your case. This is just possible due to the current state of the art both in web app development and machine learning. The culture of public AI APIs is revolutionizing the realm of artificial intelligence and its applications.

Our approach for achieving this goal is presenting a set of problems using public biomedical datasets. Those datasets are available freely at Kaggle, a platform where people make available datasets, and programmers can test their skills on those datasets; competitions are also from time to time created. We also add discussions on the topic of using models in medicine, a topic the authors have being working on for years.

We hope also to call attention to this platform, which collects a sea of freely-available datasets. As one example, our dataset for diabetes detection has 10.000 samples, even though we use only 300 samples: go back in time, a couple of years, and such public datasets would not exist so easily for machine learning testing, or any similar data science driven model.

### 1.1. Goal

Our primary objective is to demonstrate the capabilities of TensorFlow.js and advocate for its widespread adoption in the development of sophisticated machine learning models. We also hope to bring attention to JavaScript, more specifically, Angular, as a possible alternative to machine learning to traditional computer languages, namely, Python and R.

### 1.2. Contribution to the literature

We hope that our main contribution as a scientific research will be to call attention to TensorFlow.js, that is, machine learning in JavaScript. As we are going to discuss, there are several advantages of coding directly in JavaScript. Machine learning is becoming more and more present at the hands of non-experts, and TensorFlow.js may aid on that since JavaScript is one of the biggest and most used programming language, by non-experts in artificial intelligence. With TensorFlow.js, one can build an advanced neural network with little to no technical work, or actually needing to implement complex routines.

We also make available all the models we built for usage, as pretrained models, with their respective performance metrics. If the reader considers the model metrics good enough for their applications, they just need to download the model and use it.

We are actually working on a junction of those models to openAI APIs, for creating a chatbot that will call those models, without the user actually needing to call them; openAI APIs handle all the details under the hood. The results are promising: we hope to publish them soon. The fact that those models are in JavaScript made the difference: we did not need to implement a server or use another languages such as Python.

This paper can be seen as a brief tutorial on TensorFlow.js with real-world examples/applications, focused on medicine. Another way to see this paper is as a data science exercise, where we apply TensorFlow.js on medical datasets, and show what we can learn from this approach.

Therefore, we do not propose anything new, like an approach. Even though our approach to the datasets is new, using JavaScript, how we approach the problems is also new, and also the discussions we add are our own intellectual expression, those are just to add scientific context to the discussion.

### 1.3. Background

On this section, we briefly present the key concepts necessary to understand the content of this paper. Given that this paper is about bioinformatics, we anticipate a diverse audience, ranging from computer scientists to medical doctors. Therefore, it is imperative to explain these concepts. Tools such as TFJS enable individuals with minimal coding knowledge to construct their own neural networks. However, a basic understanding of these concepts is still necessary to interpret the results or to construct one’s own models. This section aims to provide an overview of these basic concepts. For further information, we recommend consulting textbooks on the subject.

#### 1.3.1. How does a machine learning model work?

A machine learning model works in two stages: first we train it, then we use it. The training is done on a dataset similar to the one it supposes to predict upon.

There are several issues that may appear. The most important one is bad dataset, as we like to say “garbage in garbage out”. One type of bad dataset is biased one. It means the dataset is not well-balanced, not well-representative; e.g., for the small dataset of diabetes, we had the care to select an equal number of samples from both classes, and randomly selected, and shuffled the dataset before training. It is important to bring to attention that biased dataset is not a machine learning problem, it is a statistics problem. If the dataset is well-sampled using statistical principles, it supposes to be good.

For eventual applications on real scenarios using our insights, please, make sure that the datasets used can represent your target population accordingly. As one example, we have no strong evidence, as far as we know, that diabetes may change significantly according to population (e.g., genetic factors), but, one should test the final model on local data before using it. This is warning is very important if you decide to use our pretrained models that we made available for each case we discuss: make sure the dataset we have used is representative for your case.

#### 1.3.2. TensorFlow.js

TensorFlow.js is a JavaScript-based library for deep learning, based on the classical TensorFlow, written in Python; you can also do simple machine learning, some simple mathematical operations with tensors and so on. We are going to build multilayer perceptrons (MPLs).

There are several reasons for using TensorFlow.js instead of Python, an imperative reason is using just one language, from the app to machine learning (i.e., JavaScript and derivatives). Another reason is that you do the calculations on the browser, no need to have high-performance servers. If your app starts to gain users, the calculation cost will not grow since each user is responsible for their calculation load. This last point is especially interesting for startups, since you can scale up without also increasing the cost. One reason for medical applications: your data never leaves the browser, it is ideal for sensitive data.

One advantage of using TensorFlow.js on the browser, instead of in the sever, is that it was well-configurated to use local NVIDIA GPUs [2]. Those computation processors are well-known for being powerful on numerical calculations, and they helped on the revolution done by deep learning.

A nice point is that they claim it is possible to transform models in both directions: TensorFlow.js to TensorFlow, and *vise versa* [1, 2, 3]. Even the manual transformation is possible since their notations are similar.

TensorFlow.js provides several ways to be used: pretrained models, CDN calls, NPM, and even downloading the model; we made our models available, you just need to download them for testing and using them on your projects. We are going to train our own model, and use it locally on a app in Angular.

#### 1.3.3. Neural networks and machine learning: supervised learning

Artificial Neural Networks, neural networks for short, are a subset of machine learning techniques, focused on numeric algorithms [9]. Different from alternatives from artificial intelligence, those algorithms are not focused on reasoning; even though non-experts using those model may think they reason [10]. Moreover, even though they are inspired by the brain, they do not replicate the dynamics of neurons with fidelity [11]. Their only goal is learning from samples, and it does not matter how; in most of the time, it is close superficially from the brain workings.

We are going to use the supervised variety: one presents a set of inputs, and expected outputs (based on human’s feed-back, a process called annotation), and the algorithm should learn, without human interference, except at annotating the dataset.

On this type of algorithms, they should learn on their own, we do not interfere with the learning process.

The separation we generally do calling them black-box models has to do with the fact that they learn, but we cannot in general explain how it works, except with metaphors how it works. See Supplementary Material for a more contextualized discussion.

One well-known warning fact about those models: we cannot ensure they will always behave as we expect: chatGPT is full of examples where its behavior is erratic compared to what we would expect from this model. What we can do, and we will do, is using performance metrics. Those are the standard to assess whether a model is minimally reliable for usage.

#### 1.3.4. Multilayer perceptron (MLP)

A Multilayer Perceptron (MLP) is an artificial neural network that has a well-defined structure, architecture.

It has an input layer (no neuron, just inputs), a hidden layer, and an output layer. Those models are widely used on regression. We are going to consider what is called logistic regression. “Logistic regression estimates the probability of an event occurring, such as voted or didn’t vote, based on a given dataset of independent variables.” IBM. Thus, our model releases a number that can be seen as a sort of probability, even if it does not have the rigour of statistical analysis; it is not a true statistical measure. This number varies from 0 to 1, and generally above 0.5 is considered as ‘yes’, and below, ‘no’; this is called threshold. This strategies is common on machine learning: every model based on classification will give this “probability” measure of a set of classes, or decision options.

We are going to consider binary classification: yes or no. This means that our model has just one output neuron; it has just two options to pick up from. On the models available on Kaggle, they use more than one hidden layers: our experience shows that just one hidden layers is generally enough, and you just need to adjust the number of neurons on the hidden layers. Thus, we always adopt the strategy of just using one hidden layer, and handpicking the hidden neurons configuration.

As one example how it looks like on TensorFlow.js for the diabetes case:

~~~
   //Input layer & hidden layer
   model.add(
     tf.layers.dense({
      //here we define the number of
      // features
      inputShape: [numOfFeatures],
      //Number of hidden neurons units: 50,
      //Activation function activation: ‘relu’,
     })
   );
//Output Layer model.add(
   tf.layers.dense(
   //just one neuron
     {
       units: 1,
       activation: ‘sigmoid’
     }
     ));
~~~

Keep in mind that the configuration (mainly number of neuron on the hidden layer may change to the other cases). In case of adaptions, use the number of hidden neurons to try out better models, with higher accuracy.

### 1.4. Organization of the work

In Methods, section 2, we talk about what is behind our results, the tools, paradigms and methodologies; our hope is that people outside bioinformatics and artificial intelligence could actually read the paper, and get insights for a possible alternative area of research. On this section, we provide also links to GitHub repositories and Google Sheets used on the simulations. We have also deployed the dataset at Kaggle, as alternative to the Google Sheets, and also as a way for Python programmers to try out their approaches if they wish so. Then we shift to results and discussion in section 3, where we talk about the simulations we did, and we use the opportunity to add our own work experience with computational biology and computational intelligence; we use graphs and tables to present our results. We finally close the results and discussion section with a summary, inside the section 3. One can find at the ending of the article our main references. We have also added a Supplementary Material at the end. There are remaining discussions that are important, but may be skipped by researchers focused on the computational part.

## 2. Methods

On this section, we present our tools, methodologies and paradigms. We have done our best to report all the important details, for a possible replication of the findings or even adaptation to a specific case. If we have forgotten anything important, please, do not hesitate to get in touch. Since we anticipate possible readers from medicine, and related medical areas, we may have been more detailed than normally those section are.

### 2.1. Training the model

The models are trained using the default routines from TensorFlow.js, more details can be found on the provided gists on GitHub on each respective subsection from this section.

This is how we set up the training configurations.

~~~
model.compile({
   optimizer: tf.train.adam(),
   loss: ‘binaryCrossentropy’,
   metrics: [‘accuracy’],
});
~~~

On a possible adaption of our reported findings, starting from this part could be a good strategy.

As optimizer for the error: Adam optimizer (or Adaptive Moment Estimation), which is a stochastic gradient descent method that is based on adaptive estimation of first-order and second-order moments, see this blog post. It is a local search, thus, the training may not always converge; alternatives are global optimizations. Except for the heart failure model on section 3.3, the two other models converged well, on all attempts; on the heart failure model, we had to make some attempts before the model would converge accordingly. This is a well-known issue when using local optimization: they are called traps.

For loss function, how we measure our error for guiding the optimizer, we use *binary Cross-entropy*, which is a binary cross-entropy metric function which uses binary tensors and returns tf.Tensor object. See this blog post. When changing the model, you may need to consider changing this function. Generally, for the kind of problem we report, this loss function is enough.

For accessing the final results, we are using accuracy, which is a ratio between what the model gets right vs. all the attempts. The ideal is 100%, and we may say that 50% is like chance, randomness. One important observation: 100% of accuracy does not mean the model does not make mistakes, just mean that using the given dataset, it did well; if the dataset is a poor representation from the case, it will fail when applied on real-scenarios.

We have also used additional performance metrics: confusion matrix, precision, F score, and recall. What is interesting: some of the those metrics are on the official documentation from TensorFlow.js, but they do not seem to work properly. All those additional metrics were calculated on a separate set of samples, that we isolated before training. We have always used 100 samples, 50 for each categories. We want to make sure the model learnt, did not memorized. We have created our own NPM package for calculating these metrics and have published it. It is available to the public.

### 2.2. Training and validation

It is a common practice in the machine learning community to validate the models splitting the dataset into learning and validation: we are using 20% for validation is 80% for training; these values are widely used.

This is how we set this configuration on TensorFlow.js: *validationSplit* : 0.2. For seeing more details, just access the gists provided. It is set on the training section, namely: *model. f it*.

Below is a sample how it looks like.

~~~
await model.fit(
features_tensor_raw, target_tensor,
{
  batchSize: 40,
  epochs: numberEpochs,
  validationSplit: 0.2,
~~~

The validation curve is used to avoid memorization, over-fitting. A model that memorizes is useless since it will have to predict outside their training dataset.

One important observation: this split was done after a preprocessing split; We split the initial dataset into 90% for training, which was split further as we presented, and 10% for performance check: we calculated the confusion matrix, precision, recall, and the F1 score. This was done to make sure the model did not memorize. Therefore, the dataset used to calculate the performance metrics was never presented to the model: we have selected 50 samples for each class (diabetes and non-diabetes). Thus, 100 samples to calculate performance metrics. This procedure was repeated for all the models we have trained.

#### 2.2.1. Confusion matrix

A very important measure on machine learning, when dealing with classification, is the confusion matrix. We are going to use it to validate binary classifiers. For instance, ‘0’ means non-diabetes, and ‘1’ diabetes. Thus, the model can have fours possible behaviors (see Figure 1). The confusion matrix is the input for calculating the remaining performance measures.

**Figure 1.**
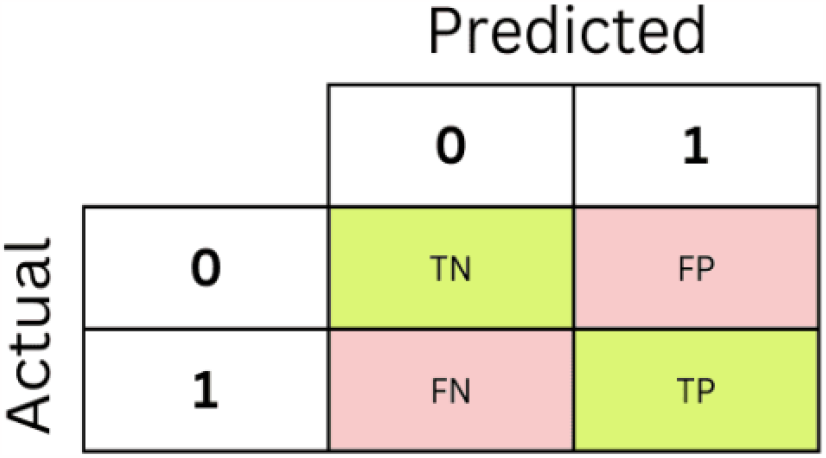
Confusion matrix. Source: adapted from KDnuggets.

### 2.3. External resources

#### 2.3.1. Pretrained models

All the models created herein are available for usage. One can find them here on GitHub. Once they are downloaded, they can be used as they are. Thus, the reader is welcome to see by themselves how the models work in action.

#### 2.3.2. Full repository

A full repository, coded in Angular using TypeScript, can be found here on GitHub. One can use those codes for replicating our findings. For specific models, we provide gists on GitHub, as so the parametrizations are also preserved, and possible specific configurations of the code. Gists are files on GitHub, which are generally used to present code snippets.

Our codes are an adaptation from [1], chapter 2; the reader can consult this book for a getting started reading tutorial. One can find here on GitHub the codes for these codes that served as starting point for our own. These details may serve someone wanting to adapt the findings to their own medical cases. One can find a sandbox here on StackBlitz, which can be forked and used for experimenting on this basic model we have used.

#### 2.3.3. Datasets

”Garbage in, Garbage out”

All the datasets were dowloaded from Kaggle. We have done the following editing, as so the datasets would fit our purposes accordingly.

1. We have sampled randomly a small number of samples from the complete dataset. In the case of diabetes detection, the sample was done manually, creating a second spreadsheet from the original. For the other cases, we have used an internal routine in TypeScript, provided inside the code;
2. We have transformed non-numeric features into numerical ones. We have also done either manually or provided a routine on TypeScript;
3. We have selected the features we wanted for each model, accordingly;
4. We have uploaded it to Google Sheets, published it and used their public spreadsheet link for uploading the dataset into our models. When the routine was local for preprocessing, we have used the full spreadsheet from Kaggle. Those details are provided on each case, on the discussions;

See that you can find the detailed preprocessing on the codes, or the final spreadsheet links on the following sections. They are all as CSV (comma-separated values) format, with heading correspondent to the feature; rows are different samples, and columns are the respective values for the feature on the heading.

##### Disclaimer

Always keep in mind: those are public datasets, we cannot guarantee those datasets are well-curated (e.g., unbiased, well-sampled and more). Nonetheless, the datasets have a score called ‘Usability’, and they are opened for comments. One can use those information to search for good datasets.

### 2.4. Diabetes detection

The complete dataset is here. The following datasets are derivations from this one, adapted accordingly to fit our purposes.

For getting the link when made available, either click on it, which could trigger an automatic download, or right-click on the link and ask to copy the link, and place it on the code; you must replace the variable ‘csvUrl’, the code was designed to adapt accordingly. You may need to change the visualization method, deciding which feature you want to plot; as alternative, just comment this method call out; the method for visualization is called ‘visualizeDataset’, just comment out the call on the ngOnInit() method, in case you want to train without visualization. In Angular, ngOnInit() is a hook, in this case, make sure something will happen just after the app is started, see [5] for more.

#### Importat

For the upcoming sections: keep in mind that features are the input to the model, it does not count the output (e.g., diabetes detection).

##### 2.4.1. 6-feature model

The code used to train the model can be found here as a gist, the TypesScript file. Since we are concentrating on the logic of the Angular app, we are not changing the remaining component’s files. One can find in case of interest a complete repository here on GitHub, with interface from an Udemy course we have lectured. For the case of diabetes, we have at the moment a working app on Heroku, deployed for presenting the concept.

The spreadsheet link is here. As alternative, the smaller dataset we used is also on Kaggle here, we have deployed it for your convenience.

##### 2.4.2. 1-feature model

The spreadsheet link is here. We have also deployed this dataset to Kaggle as an alternative to work with this dataset.

##### 2.4.3. 3-feature model

The spreadsheet link is here. You can find here a gist, which includes the link. You can also find the dataset at Kaggle.

### 2.5. Predicting In-Hospital Surgery Complication

The spreadsheet used is here; you can also download this dataset from Kaggle. The original dataset is here on Kaggle.

This dataset is sadly not very clear on details, we have used a dictionary provided by an user, it can be found here. We used this dictionary to understand what each feature meant, as so we could build our model according to what we wanted to study: complications on surgery based on physiological measures. A gist for the TypeScript file is here on GitHub.

### 2.6. Heart failure prediction model

A gist on GitHub is here. The full dataset is here on Kaggle; the spreadsheet used on the code is here. The coded provided, different from the one for diabetes, allows to actually change the sample size from the full dataset. Just adjust *number o f samples* in *dataset to array* method. It may be useful in case you want to run the model with more samples, or make simulations using another dataset.

### 2.7. NPM respository

In addition to use NPM respositories, such as TensorFlow.js, we have also created our own. We were unable to find a package with the basic metrics for model validation. We created one and deployed it as a NPM package.

It is possible to install it on your own project by running on the terminal:

~~~
npm i tfjs-metrics
~~~

Learn more on the package page on NPM we have created and deployed. The performance metrics we used were calculated using the routines on this package, which can be used by anyone coding in TensorFlow.js focused on Angular (TypeScript).

## 3 Results and Discussion

Below, Table 1, is a table that summarizes all the results, using basic metrics for assessing the performance of those models after training is completed. The upcoming section will detail these numbers.

**Table 1.** Performance metrics for each model.

| model | Confusion matrix |  |  |  | Performance metrics |  |  |
| --- | --- | --- | --- | --- | --- | --- | --- |
|  | TN | FN | FP | TP | recall | precision | F1 score |
| 6-feature diabetes | 43 | 7 | 5 | 45 | 0.89 | 0.86 | 0.87 |
| 1-feature diabetes | 42 | 9 | 8 | 41 | 0.82 | 0.84 | 0.83 |
| 3-feature diabetes | 30 | 8 | 20 | 42 | 0.78 | 0.6 | 0.68 |
| Surgical Complication model | 50 | 0 | 0 | 50 | 1 | 1 | 1 |
| Heart Failure model | 35 | 4 | 15 | 46 | 0.89 | 0.7 | 0.78 |

We are going to discuss three different group of models: diabetes on section 3.1, heart failure on section 3.3, and surgical complications on section 3.2. Those are all binary classification tasks: ‘0’ (no) or ‘1’ (yes). For the reader interested, we have a list of datasets for binary classification tasks here, as so you can adapt our discussions for the case of your interest.

For the diabetes model, we have created three different models to discuss how features can influence the model: the 6-feature model has six different features, medical measurement. The 3-features uses three features that are easy to have, that is age, gender and BMI. The 1-feature model uses the most important one: HbA1c level. In fact, we have showed using this same dataset that gender has an effect that is statistically significant on diabetes [12]: it is more likely in men. We have also explored this dataset on our other work [12].

### 3.1. Diabetes detection

Diabetes is an interesting case for our discussion, it has been widely explored from both perspectives, as machine learning based models ([13, 14, 15]), and white box models [16].

On this section, we shall consider three possible models for diabetes detection using the same dataset, and model. The first model has six features, the second just one, and the last one has three features. The features are chosen from the same set of features.

Always keep in mind that our goal is not comparing results or competing with third-party results presented, they are presented for enriching the discussions. Our goal here is showcasing TensorFlow.js, and supporting on spreading the word regarding this tools for creating advanced machine learning models. Years ago, those models would require either a strong expertise in programming and machine learning, or/and an expensive-paid software (e.g., Matlab).

#### 3.1.1. model 1: predicting with six features

We should always start with the simplest model, nonetheless, we shall start with the almost-full feature model. We are going to use the following features (Table 2), six out of seven available features.

**Table 2.**
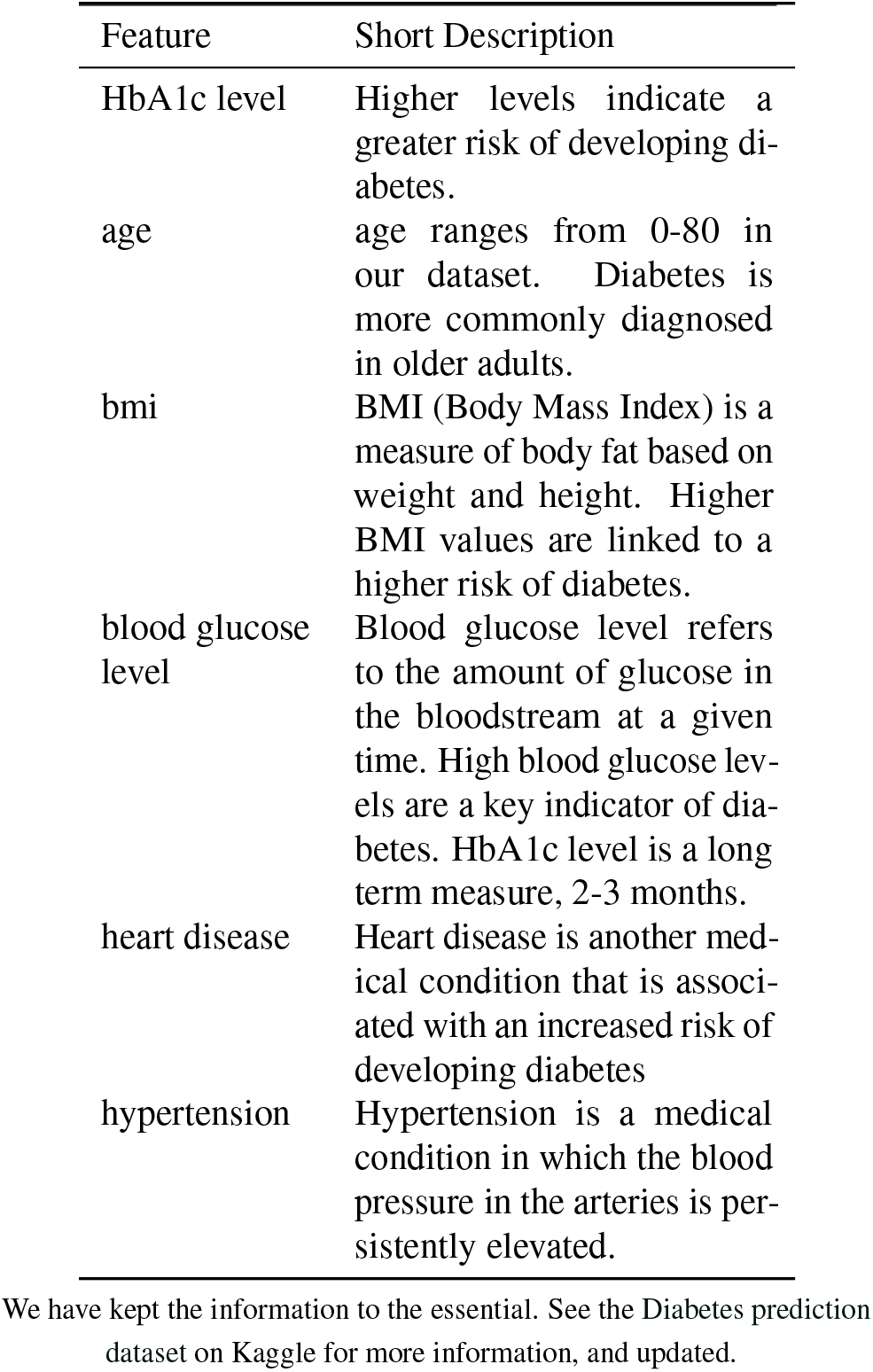
Features used to train the model with six features for diabetes detection.

| Feature | Short Description |
| --- | --- |
| HbA1c level | Higher levels indicate a greater risk of developing diabetes. |
| age | age ranges from 0-80 in our dataset. Diabetes is more commonly diagnosed in older adults. |
| bmi | BMI (Body Mass Index) is a measure of body fat based on weight and height. Higher BMI values are linked to a higher risk of diabetes. |
| blood glucose level | Blood glucose level refers to the amount of glucose in the bloodstream at a given time. High blood glucose levels are a key indicator of diabetes. HbA1c level is a long term measure, 2-3 months. |
| heart disease | Heart disease is another medical condition that is associated with an increased risk of developing diabetes |
| hypertension | Hypertension is a medical condition in which the blood pressure in the arteries is persistently elevated. |

We have kept the information to the essential. See the Diabetes prediction dataset on Kaggle for more information, and updated.

The best results on the codes available alongside the dataset, using TensorFlow in Python, they have used all the features: seven at total. See here a list of notebooks modeling this same dataset on Kaggle using Python. The nice feature of this dataset is that they have attached codes, created by the users, and publicly available, and editable. TensorFlow in Python and TensorFlow.js have similar notations: we have explored those codes for starting our models when necessary. We have removed the following feature: smoking history. We got essentially the same result, which means that smoking history, at least for this group, does effect diabetes significantly. In fact, machine learning is also about statistical analysis. We tested the hypothesis of smoking being connected to diabetes: we found that there is a connection. See Supplementary Material; see also [17]. Thus, a 7-feature model may be interesting to build. One way to explain the success of the model without smoking history is that smoking may cause/influence the other features, therefore, they are correlated.

The best model on Kaggle arrived to 97% of accuracy, see here, we have arrived to 92% of accuracy (Figure 2); sadly, we were unable to see the validation results for making sure they had no overfitting, for the notebooks on Kaggle attached to the dataset. This extra accuracy presented by the developer at Kaggle was done in addition of using an extra feature by applying Random Forest Regressor (i.e., a different technique), see here. Since the training process can give out different results depending on the starting point and this high accuracy was achieve with an alternative approach, we cannot say for sure smoking accounts for this 5% of lost accuracy. Those third-party results were mentioned for the sake of information, it is not our goal here to compare different approaches on machine learning for logistic regression, as generally data scientists call this problem. We have also not checked carefully the results on those notebooks on Kaggle, for making sure they did not take shortcuts. Here on this notebook they have also found 97% for a model similar to ours, a Multilayer Perceptron (MLP).

**Figure 2.**
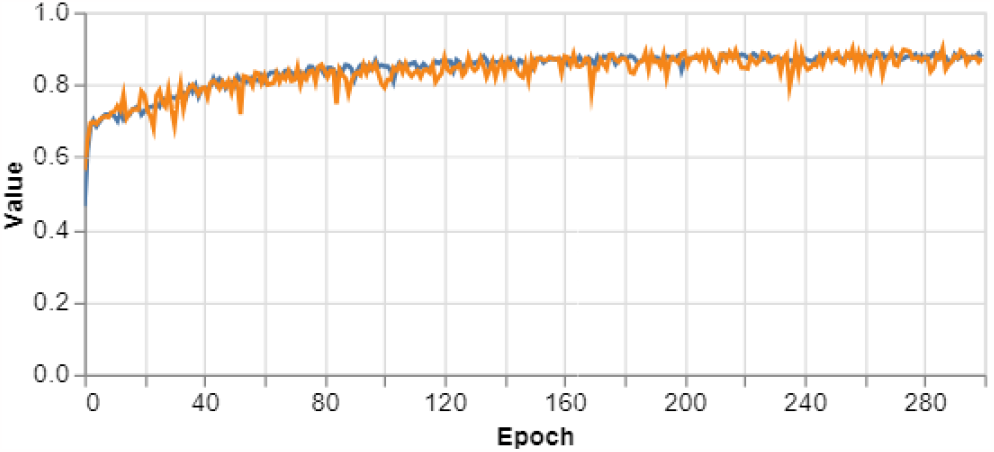
Accuracy for training with six features. final result, about: 92% for training and 85% for validation. Training in blue and validation in orange.

Below is the training curve (Figure 3). One can see that both training (in blue) and validation (in orange) curves go downwards, and remain at low values. This is a sign that the model actually learned and was able to generalize. No overfitting happened, based on those graphs. Additionally, one can check further metrics (see Table 3 and the following metrics calculated using this confusion matrix).

**Table 3.** Confusion matrix for the 6-feature model. Legend: TP - true positive; FP - false positive; FN - false negative; TN - true negative. The row label represents the true class, whereas the column label is the model prediction.

|  | non diabetes | diabetes |
| --- | --- | --- |
| non diabetes | 43 (TN) | 7 (FP) |
| diabetes | 5 (FN) | 45 (TP) |

**Table 4.** Confusion matrix for the 1-feature model. Legend: TP - true positive; FP - false positive; FN - false negative; TN - true negative. The row label represents the true class, whereas the column label is the model prediction.

|  | non diabetes | diabetes |
| --- | --- | --- |
| no diabetes | 42 (TN) | 8 (FP) |
| diabetes | 9 (FN) | 41 (TN) |

**Table 5.** Confusion matrix for the 3-feature model. Legend: TP - true positive; FP - false positive; FN - false negative; TN - true negative. The row label represents the true class, whereas the column label is the model prediction.

|  | non diabetes | diabetes |
| --- | --- | --- |
| non diabetes | 30 (TN) | 20 (FP) |
| diabetes | 8 (FN) | 42 (TP) |

**Figure 3.**
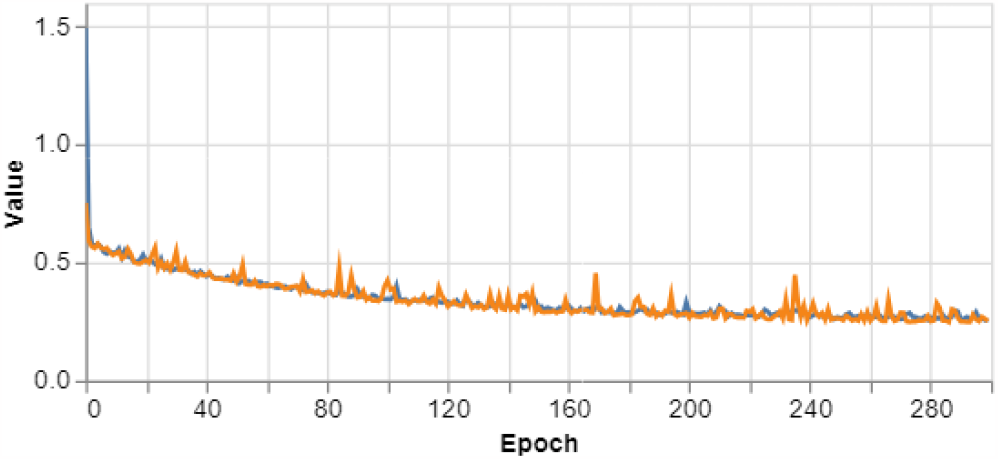
Training and validation curves for the 6-feature diabetes detection model. Training in blue and validation in orange.

One point that we should bear in mind: we have not used all the dataset. The complete dataset of diabetes detection has 10.000 samples: we have used just 300 samples (equally balanced between diabetes and no-diabetes, and randomly samples from this pool); a total of 600 samples. It shows how the signals on the dataset, which the model was able to learn, is strong; it also converged without any instability, which is a good sign. One can test a model with the full dataset. Since we are using the browser, uploading the dataset locally for training, it may be slow this uploading process of actually using the complete dataset; we see no reason based on those graphs to keep moving around 10.000 samples of data.

Another point is which features to use, since we have decided not to use them all, it may raise this question. In real scenarios, it may be the case that the person doing the model does not have all the features; or even when applying the model, the user does not have all the features. We are going to consider two extra scenarios in the upcoming sections: choosing the most important one (namely, HbA1c levels) on section 3.1.2, and the three easiest ones to have access to (namely, BMI, gender and age), on section 3.1.3.

From the user’s perspective, one can build different models with different features, and this change is made according to how much information the user has. Of course, the prediction accuracy will change accordingly. As we are going to see, the accuracy will drop to about 70% in both cases where we have not used all the 6 features.

We have recently created and tested a chabot using openAI APIs that can shift between models, based on chat messages. We tested for the 6-feature model alongside models for image, and it worked; we are planning to publish the results soon. Thus, all those models could be automatically used by a chat-bot (a conversational AI) according to how much information the user provides, under the hood, the user will not have to change any model manually.

One good question is why just one, or even three features, can achieve 70%, leaving the remaining with 20%. You can find an explanation on Figure 8 (correlation matrix): the features are correlated, which means they do not convey pure information.

From a developer’s perspective, one can build the model based on the features available, or measure them accordingly to their power of prediction. We are going to consider a model with just one feature, the one that predicts the most. We are thinking, as a hypothetical scenario, one where one must decides which features to invest on to measure on several patients.

The performance metrics are for the 6-feature model are: F1-score, 0.87; precision, 0.86; recall 0.89. The execution time was less then 5 minutes, on the browser. All the upcoming models had similar execution time.

The model trained, and ready to use, can be found here on GitHub. One example on how to upload a pretrained model to Angular, using this JSON file, is here on GitHub.

One interesting discussion is how to use this model when we do not have a big dataset; we showed that the model converges even for 300 samples, which may be big if we are using a small town as target for our model, or creating a prototype. A quick search on the internet points out that people are considering transfer learning in regression: it is generally applied to images [18]. We are considering, given we have access to those data in the future, using this model in Brazil. Our current setback is actually having access to this data as we have on Kaggle, publicly and easily accessible. Our concern is that this dataset may be biased; biased dataset is a well-known issue on machine learning, they contaminate the learning process, and the machine learning will make mistakes when confronted with categories misrepresented on the dataset used to train. We have a guess that population variations (e.g., genetics, diets) may effect how those features predict diabetes.

Regarding the dataset, our hypothesis is: we cannot trust on this dataset 100% since it is concentrated on USA. We may need, or anyone considering actually using this model locally, make some adjustments to make sure the model is not biased towards the USA population, and their biological, cultural peculiarities.

#### 3.1.2. model 2: predicting with just one feature

We consider just one feature, namely, HbA1c levels, we create three regions when this choice is applied to our dataset (Figure 4): two has diabetes diagnosis well-defined, whereas the third one is a grey area.

**Figure 4.**
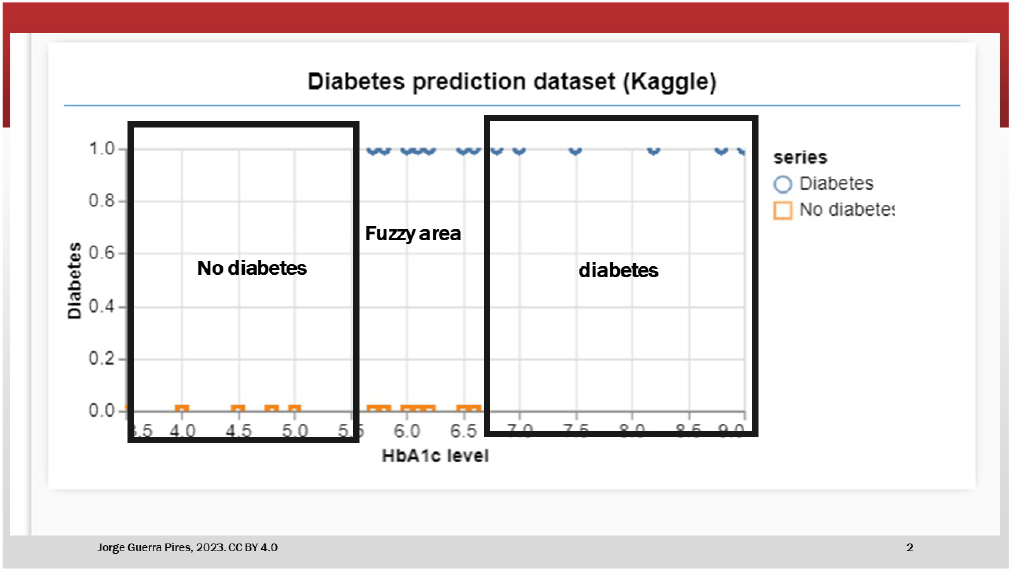
Predicting with just one feature. We have three areas: two are well-limited by HbA1c levels, whereas we have a grey area. This area was created considering what is normally accepted on the medical literature, just an approximation.

Regarding this grey area, it is interesting to mention that even though we shall almost completely shed light on it, mis-diagnosis of diabetes due to other medical conditions is not uncommon (e.g., [19]). As one example, liver disease can also alter HbA1c levels, and we have no information on that on our dataset, as a feature, whether the patient has liver disease, a richer list created by chatGPT here. Liver dysfunction or certain liver diseases, such as cirrhosis, can affect the production and breakdown of hemoglobin. This can lead to changes in HbA1c levels [20], potentially leading to misleading results in diabetes diagnosis.

This regions are based on numbers we can find online regarding how to interpret this coefficient; we also had a big help from chatGPT.

Figure 5 illustrates that the accuracy falls to 72%, the removed features account for about 20% of the 6-feature model accuracy. As said before, it does not mean this is a generalization, that this ratio between lost accuracy and the one the model has will always hold true: if someone has liver disease, as highlighted previously. Unknown medical information from the patient may mislead the model, and this feature is not present at the model as so it could actually learn to avoid this diagnosis trap.

**Figure 5.**
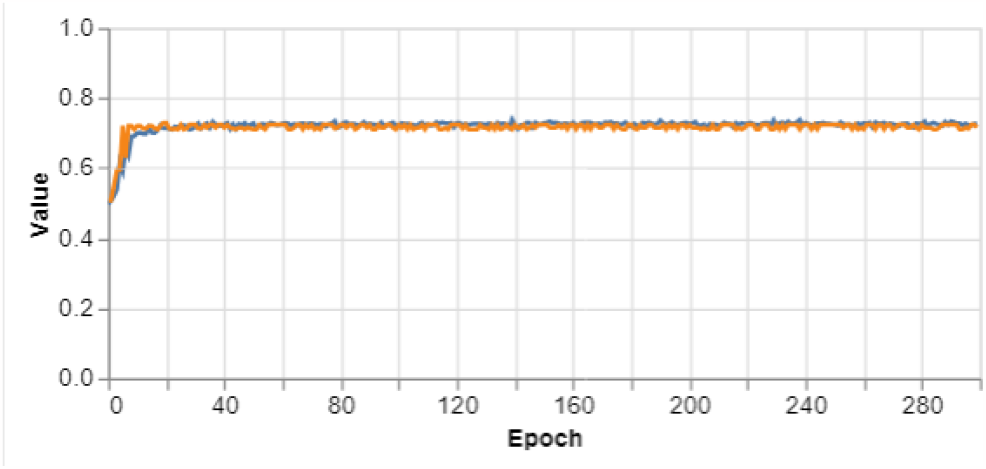
Accuracy for one-feature model, training (blue) and validation (orange). Final: training accuracy, 72%; validation 74%

A scientific question worth mentioning is how we decided to use just this feature, which has 70% alone of accuracy: just this feature is better than random guess. See discussion based on Daniel Kahneman and colleagues on Supplementary Material. Would this model be better than an expert? to which extent? good question! As they have noticed, in some scenarios, a random model was better than human; it was observed in clinical practices [21]. For image, it was shown that a transfer learning model on medical images was better than human experts [22].

One way to look at it is by rationale. Glucose is the number one factor that appears on white-box models [16], it is not by change; those models aim at explaining datasets by actually understanding the inner dynamics from the biological system, different from machine learning, which is just focused on somehow learning and predicting. However, the fact that your glucose is high for a couple of days does not make you diabetic (short-term factor, see Figure 6); but still something to pay attention to, your glucose dynamical system should be able to respond daily. HbA1c is measured in months (long-term factor), thus, it is a measure of high-blood glucose level persistence. It may shed light on why this factor is so powerful on predicting. As we are going to see in the next model, glucose alone account for about 20% of the signal for predicting diabetes, using correlation as indicator.

**Figure 6.**
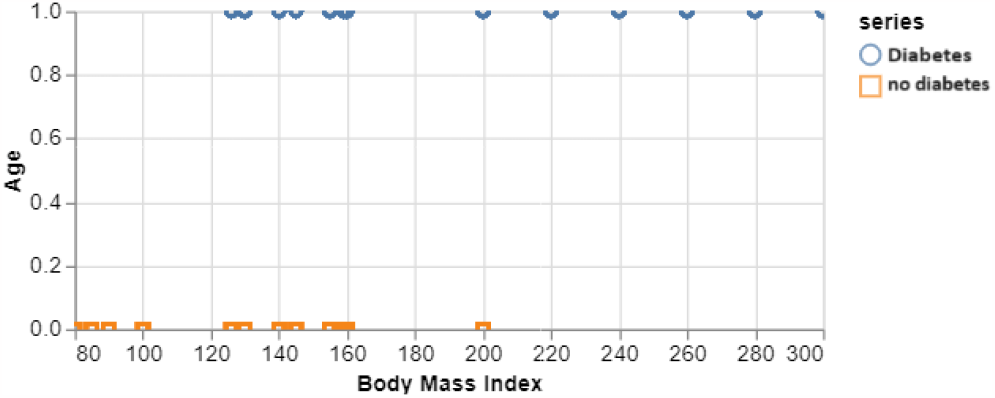
Glucose levels vs diabetes. Note. this graph shows a “simultaneous measure”, it is not a cause-effect measure, the fact that you have (uncontrolled) diabetes entails high glucose, but high glucose does not necessarily mean diabetes, though most likely. As we are going to see, glucose accounts for about 20% of the accuracy, not 100%.

**Figure 7.**
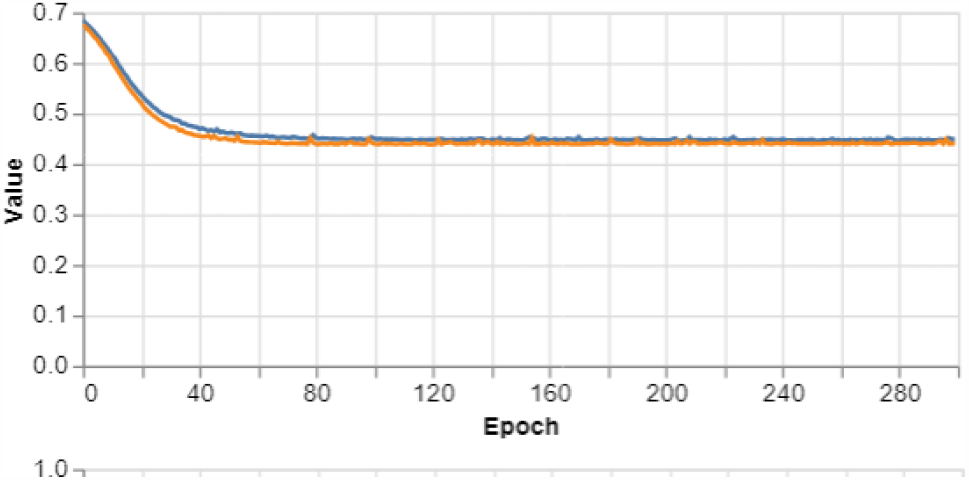
Learning curves for training (blue) and validation (orange) for the one-feature model.

**Figure 8.**
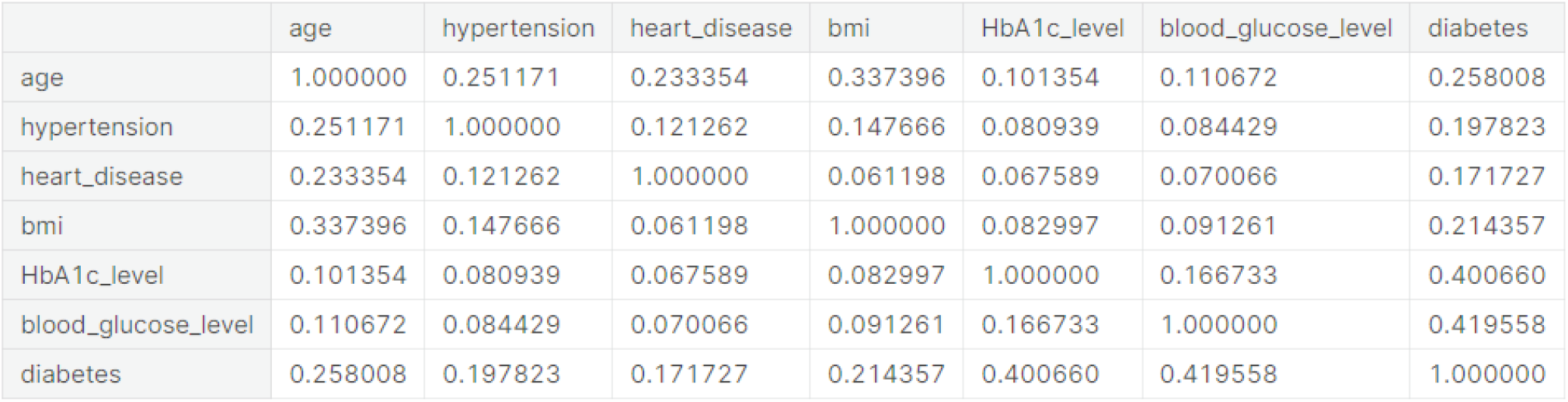
Correlation between the diabetes detection features. Source: Notebook on Kaggle

The remaining performance measurements: F1 score, 0.83; precision, 0.84; recall, 0.82.

Another way to look at it is by correlation. Correlation measures the connection between two variables: it can be positive ( from 0 to 1) or negative (from -1 to 0). Even though correlation is a linear-relation indicator, if used with attention, it can help us even on nonlinear cases; which is the case for most problems in medicine. However, one should always pay attention to the fact that it measures linear relationship, and can misdirect on nonlinear cases.

As we can see from the diabetes column (Figure 8), the two highly correlated with diabetes detection are glucose-related. Thus, the best way to diagnosis diabetes is by glucose-related indicators. Diabetes is well-studied, therefore it is no surprise, nonetheless, for less studied cases, when you apply those models, you may want to get to know the inner dynamics before pouring numbers on the model, that is the take home message.

The pretrained model can be found here on GitHub as a JSON file.

#### 3.1.3. model 3: predicting with age, gender and body mass index

In Figure 9, we have a look at how body mass index (BMI) and age may influence diabetes; keep in mind we have used it on our 6-feature model, we are just narrowing it down to easy-to-use features, namely, body mass index (BMI), gender and age. Those are three features that anyone has access to.

**Figure 9.**
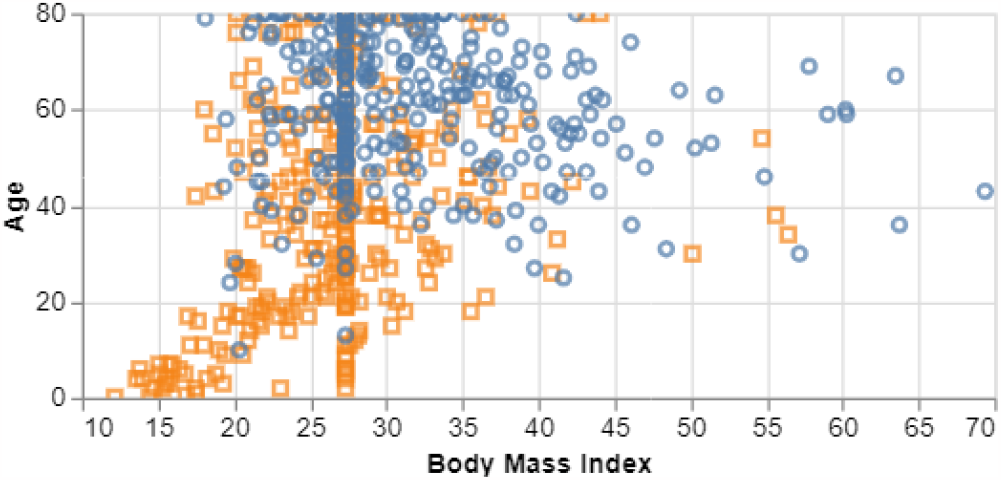
Body mass index vs. age. Normal BMI ranges from 18.5 to 24.9. Diabetic in blue, and non-diabetes in orange.

It is possible to infer, in blue, that diabetes is present at high BMI, and very rare at low BMI. Diabetes presents itself mainly as high-blood sugar, known as “sweet urine”; the body tries to get rid of the extra glucose in the urine, and this can cause a sweet smell. Obesity (high BMI) can cause difficulties on cells’ glucose receptors, which cases what is well-known as diabetes type II, which is generally reversible based on change of habits [23].

Figure 8 illustrates a strong correlation between age and BMI with diabetes, with each factor accounting for approximately 20% of the variance. An intuitive way to look at correlation is “the correlation between two variables is their percentage of shared determinants.” [24]

Figure 10 illustrates almost the same accuracy for the 1-feature model (Figure 5); Table 1 illustrates that the 1-feature model can be even better at some metrics. This observation can be explained on a speculation level by observing that HbA1c levels are correlated with both features we have used, 10% with age.

**Figure 10.**
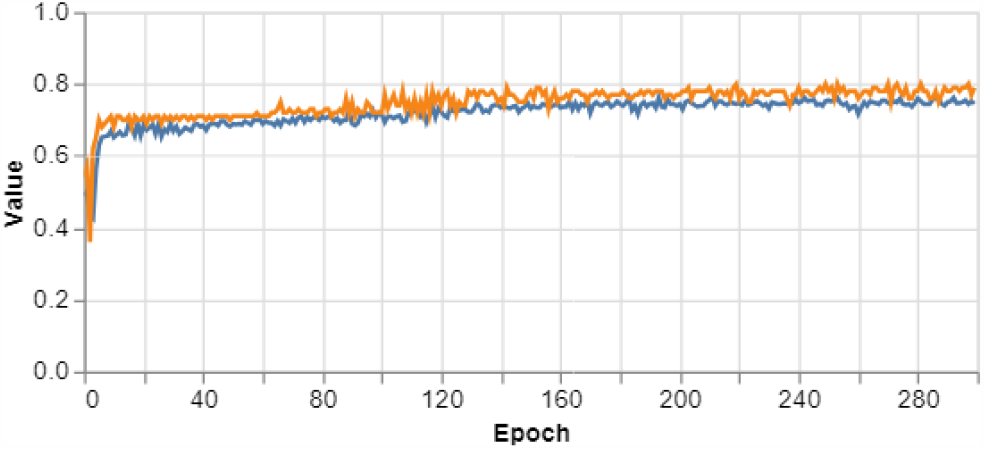
Accuracy 3-feature model. Training (blue curve); validation (orange curve)

The remaining performance measurements: F1 score, 0.68; precision, 0.6; recall, 0.78.

You can find the final model here on GitHub as JSON.

### 3.2. Predicting Surgical In-Hospital Complication

The dataset for surgical complications has 25 features that could lead to surgical complications, we are going to be even more selective on what to consider on our model. We have a hypothesis on what could lead to complications on surgery (Table 6).

**Table 6.** Features used to train the model for surgical complications.

| Feature | Short Description |
| --- | --- |
| baseline diabetes | patient has diabetes. |
| BMI | body mass index. This a measure of body weight related medical conditions, such as obesity and undernutrition |
| Age | patient age. We generally know that age may influence the final result |
| Gender | patient gender |

We have used an external dictionary to make sense of the features, see here.

**Table 7.** Confusion matrix for the surgical complications model. Legend: TP - true positive; FP - false positive; FN - false negative; TN - true negative. The row label represents the true class, whereas the column label is the model prediction.

|  | non complications | complications |
| --- | --- | --- |
| non-complications | 50 (TN) | 0 (FP) |
| complications | 0 (FN) | 50 (TP) |

The model arrived at about 100% of accuracy. Figure 12 shows the loss function arriving to almost 0 (measure of the model’s mistakes, compare with Figure 13). Also, Table 1 shows that the performance metrics are at their maximum values, that is, 1.

**Figure 11.**
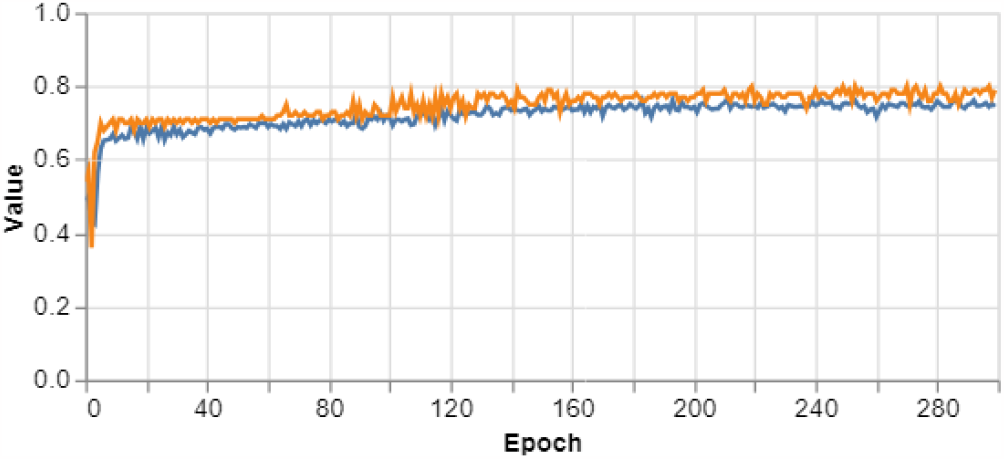
Learning curves for training (blue curve) and validation (orange curve).

**Figure 12.**
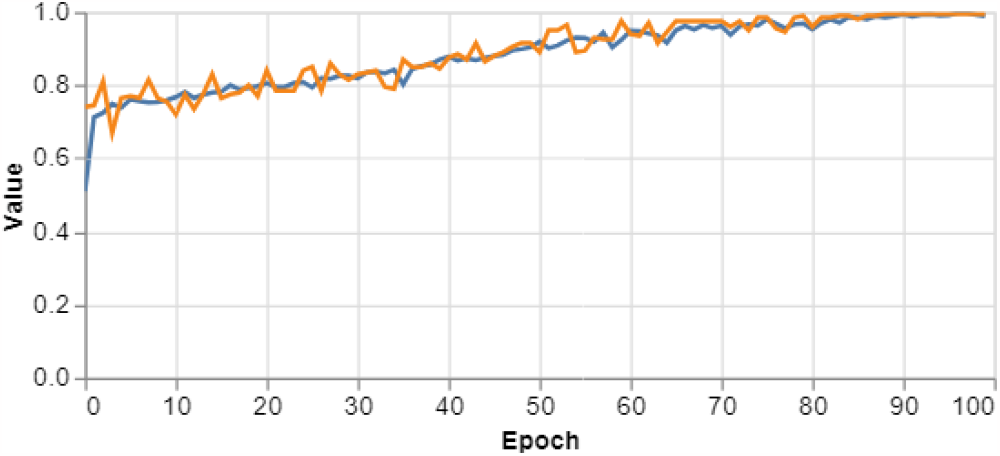
Accuracy for the surgical model. Training in blue and validation in orange. Final: training 98% and accuracy 99%

**Figure 13.**
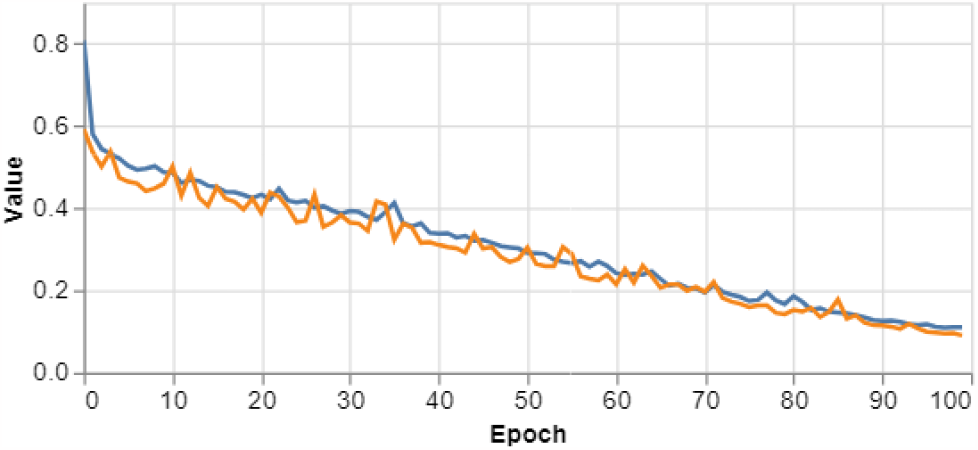
Loss function for the surgical model. Training in blue and validation in orange

The first caution we would have when seeing those results is overfitting (i.e., the model memorized the training dataset, but did not actually generalized). The validation curve also converged, which is an imperative measure to avoid overfitting, and aiming at generalization.

The remaining performance measurements: F1 score, 1; precision, 1; recall, 1.

The model was made available at ready to use as JSON here on GitHub.

#### Disclaimer

for using this model on real scenarios, please, make sure the dataset is not biased against your aimed population. We cannot guarantee the dataset used is well-curated, and unbiased.

### 3.3. Heart Failure Prediction

Cardiovascular diseases (CVDs) are the number 1 cause of death globally, taking an estimated 17.9 million lives each year, which accounts for 31% of all deaths worldwide. Four out of 5 CVD deaths are due to heart attacks and strokes, and one-third of these deaths occur prematurely in people under 70 years of age. Heart failure is a common event caused by CVDs and this dataset contains 11 features that can be used to predict a possible heart disease. People with cardiovascular disease or who are at high cardiovascular risk (due to the presence of one or more risk factors such as hypertension, diabetes, hyperlipidaemia or already established disease) need early detection and management wherein a machine learning model can be of great help. dataset description

We have selected a couple of features (Table 8), and our model achieved an accuracy of 70% (Figure 14). One user from Kaggle claims an accuracy of 90%, see here the notebook. Thus, adding all the features we may arrive to this result. This is a scientific detail we may investigate in the future.

**Table 8.**
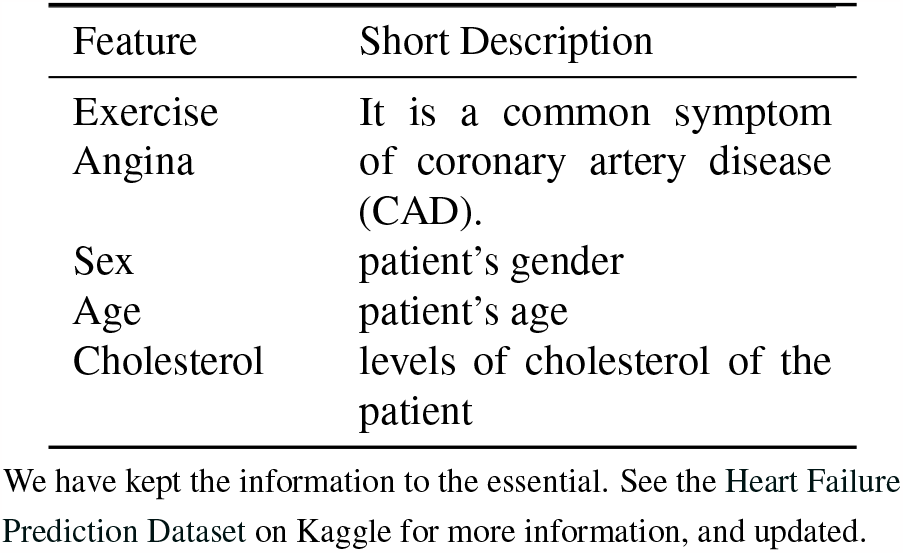
Features used to train the heart failure prediction model.

**Table 9.** Confusion matrix for the heart failure model. Legend: TP - true positive; FP - false positive; FN - false negative; TN - true negative. The row label represents the true class, whereas the column label is the model prediction.

|  | non heart failure | heart failure |
| --- | --- | --- |
| non heart failure | 35 (TN) | 15 (FP) |
| heart failure | 4 (FN) | 46 (TP) |

**Figure 14.**
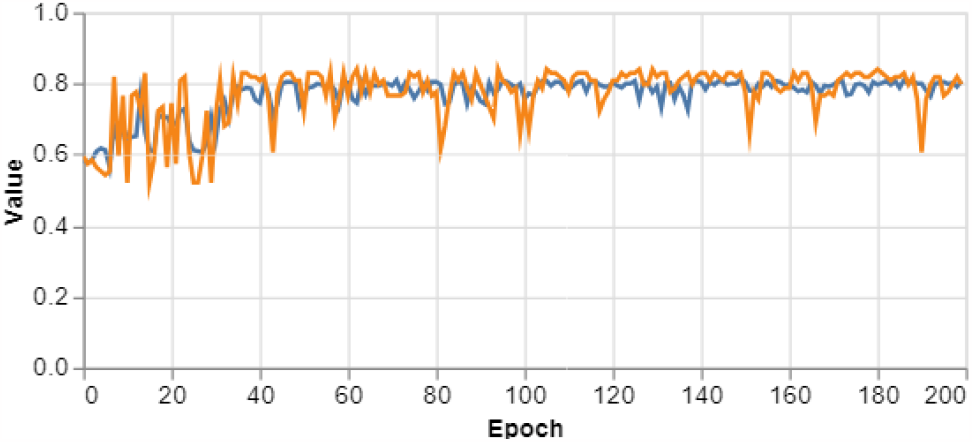
Accuracy for our heart failure prediction. final. training 80% and validation 79%

**Figure 15.**
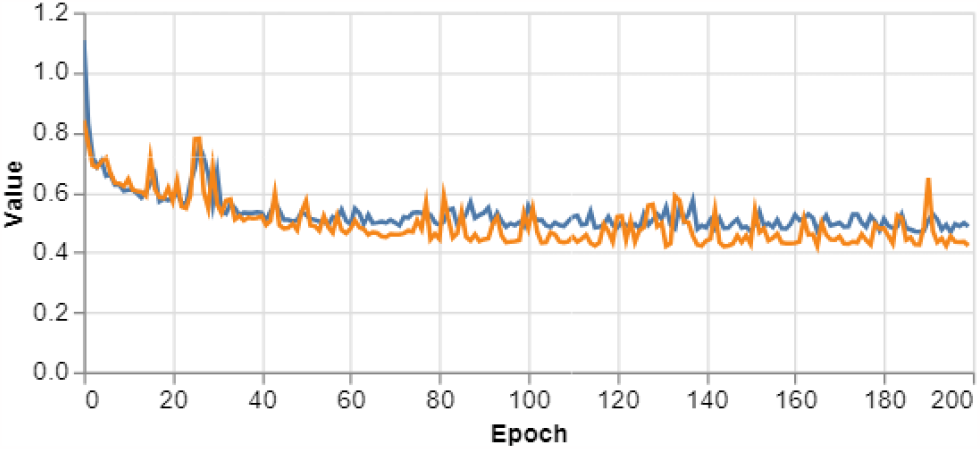
Training curves for the heart failure prediction model

**Figure 16.**
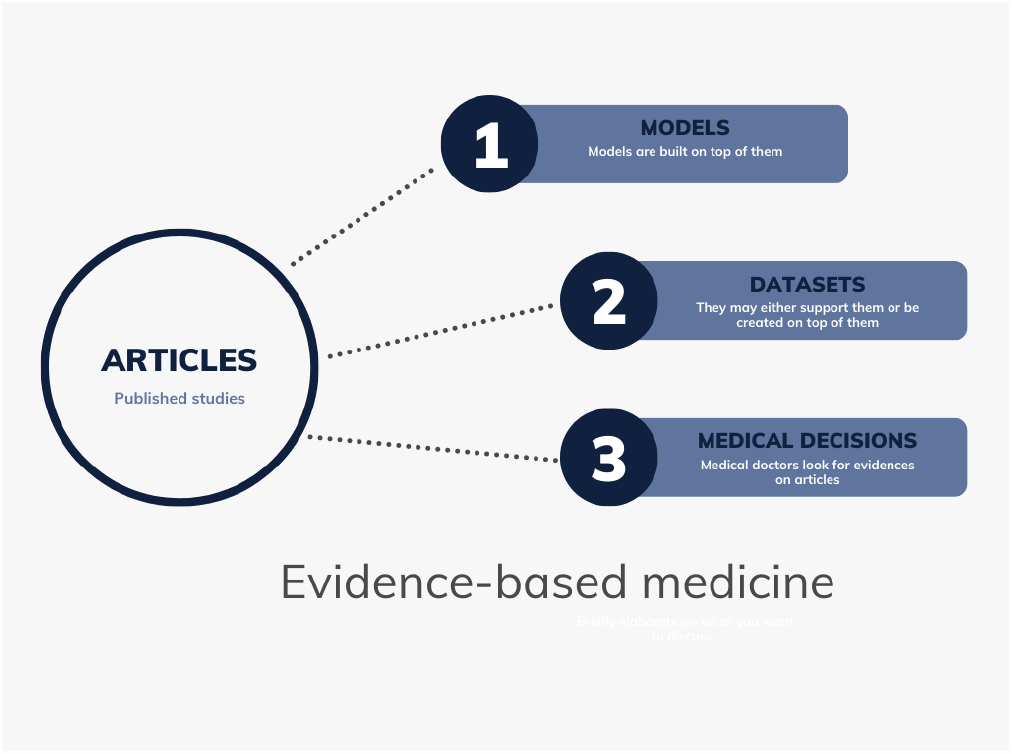
Even though medical doctors and bioinformatician may disagree on how they see evidence-based medicine, they agree on scientific papers as source of knowledge for decision making processes

**Figure 17.**
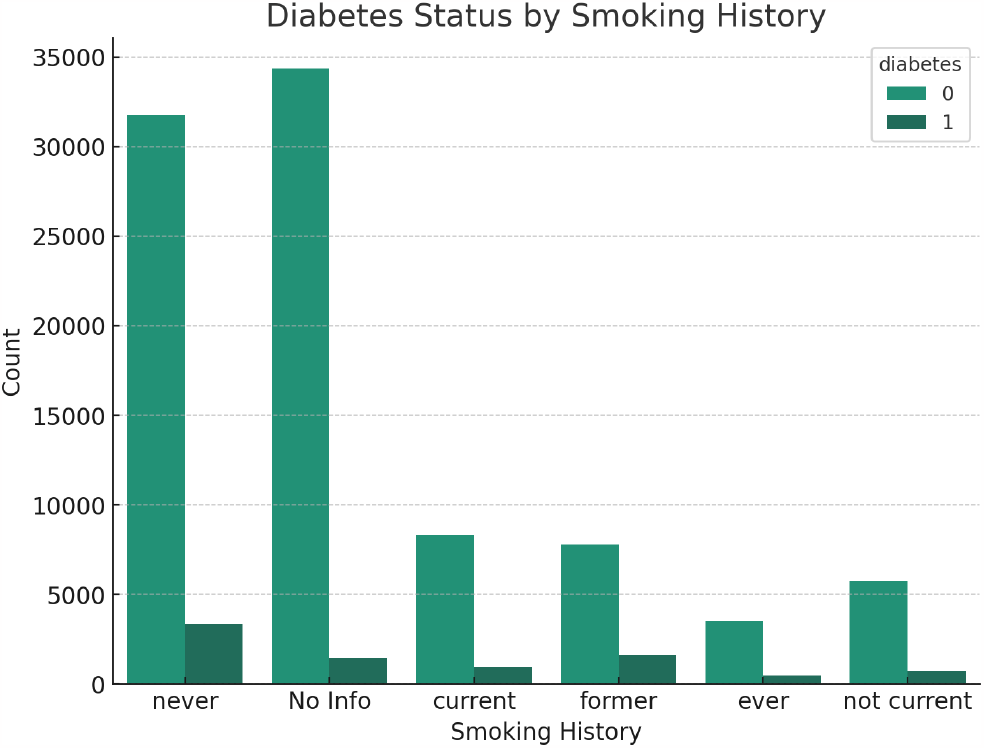
History for smoking history for diabetic patients

The remaining performance measurement: F1 score, 0.78; precision, 0.7; recall, 0.89.

The final model, ready to use as pretrained model, is available at GitHub.

#### Alert

it seems there is wrong title for this dataset according to one user. It seems the dataset predicts coronary heart disease. It does not change our results. We assume that when you use those models on a dataset in a real-case, you know the dataset enough.

### 3.4. In summary

Several examples were developed using TensorFlow.js as a machine learning platform in this manuscript. Even though Python and R are dominant on the moment, JavaScript and derivatives are growing fast, offering basically the same performance, and some extra features associated with JavaScript; JavaScript now even offers desktop options for coding, it is a full stack language (MEAN Stack) [25]: it is possible to build back and frontend without without needing to shift languages. Moreover, Kaggle, the public platform from where we downloaded our datasets, offers a huge amount of datasets for biomedical cases, thus, the reader can easily test what was discussed, using the same codes, with minor chances, on any case they may be interested in. After using TensorFlow.js for about three years, and after have used previous options such as Matlab, we are optimistic that this library developed in JavaScript has what it is needed to be a machine learning option for biomedical problems. They offer unique advantages, such as the data never leaves the browser; and the calculation load stays locally on the browser, no need to pay for high-performance servers. Angular, coded in TypeScript, a derivative from JavaScript, has an active community, releasing updates every six month. The possibilities are unlimited, and we believe that it is a nice option for researchers aiming at web applications, especially, focused on medicine.

## Data Availability

All data produced are available online at Kaggle (Machine Learning and Data Science Community)

https://www.kaggle.com/datasets/jorgeguerrapires/predicting-diabetes-with-6-features/data

https://www.kaggle.com/datasets/jorgeguerrapires/1-feature-model-for-diabetes-detection

https://www.kaggle.com/datasets/jorgeguerrapires/dataset-surgical-binary-classification-reduced

http://robodoc.ideacodinglab.com/#/tools/diabetes

## 4 Supplementary Material

We have decided to keep the main paper focused on building neural models using TensorFlow.js. Nonetheless, those models are applied to medicine, and we want to also add to this discussion. The final user are medical doctors, either as user or even as researcher. As one example, recently we have integrated the 6-feature diabetes model to openAI APIs creating a smart chatbot: the bot can extract the parameters from a text entered by the user, call the model, interpret the prediction, and reply back, all done as text, as a human-like conversation. We are planning to also integrate the remaining models we have discussed. This section will extend the discussion, but, focused on the big picture: how those models are used, and the context there are in. We hope to show the “big picture”, instead of just focusing on punctual results.

### 4.1. Evidence-based medicine by machine learning

Science is all about evidences. As so, some areas are classified as primary, and others as secondary, being empirical evidences as the separator. Primary areas deal with evidence at first hand (e.g., biology and physics); whereas secondary makes use of them (e.g., medicine and bioinformatics). Both medicine and bioinformatics have been using the name evidence-based medicine, they agree on the term evidence (e.g., scientific articles), but they see the term slightly different; bioinformatics also use datasets as evidences.

One may say that medicine stays on the scientific papers as evidences, whereas bioinformatics goes beyond, we build models, that can in sequence be used to take decisions. Our models are based on evidences, and can also be seen as evidence.

Our invitation is that medical doctors go beyond papers, and start to use models. We have showed and discussed a group of models known as *machine learning*. They can be used to assist on the decision-making process.

One advantage of models, compared to “bear-head model” is that they can take into account many more features and samples at once, and they can find hidden patterns that humans can not, or may miss a lot. Machine learning is well-known to be good at finding patterns that humans can not see on datasets.

### 4.2. Models in medicine: mechanical vs. clinical judgement

Mechanical judgments are when algorithms take decisions, whereas clinical judgment is when humans take decisions, concept introduced by Daniel Kahneman and colleagues [24].

We build models, as so they can support us on taking decisions, based on facts, on evidences; it is what we from bioinformatics call evidence-based medicine. Even if you are not aware, humans also consider facts, data, evidences, when deciding; medical doctors are also considering evidence-based medicine, nonetheless, focusing just on facts rather than models. The difference is that humans can not use massive datasets, integrative approaches and more. Another difference is: we can not write down clearly how we decided, even though we may think we can. We are contaminated by biases and noise, as highlighted [24]. Aware of or not, we cannot handle multiple sources of information, in our models called features; we simplify complex problems, and decide upon those reality-drafts. We create simplified versions of the problem, as so we can handle them.

This concept is valuable for us: we have actually considered situations where algorithms may actually compete with humans; of course, it is becoming more and more present. However, medicine is an area that has been resistant, even though the models are promising; models have been used on the industry for decades for supporting decision making, called *operations research*. There are several unsolved questions, and one is about accountability when algorithms make mistakes. Those discussions ought to happen at some point if we want to have more models on medicine in the future.

Daniel Kahneman and colleagues [24] brings to attention that even randomly initiated models may be better than humans, in certain scenarios.

> “In one of the three samples, 77% of the ten thousand randomly weighted linear models did better than the human experts. In the other two samples, 100% of the random models outperformed the humans.” [24]

Of course, they are considering specific scenarios, and care is necessary before big generalizations. The important message is: models, even simple ones, can replace humans. The point is: when talking about complex models, big models, integrative models, humans do not have a chance. Nonetheless, even for simple models, humans can be replaced by algorithms; and we should not neglect it based on human’s feelings of being left out.

One possible benefit of applying models to medicine is eliminating repetitive tasks, and possibly allowing the doctor to actually concern about the patient. Most of the routine tasks done by medical doctors can be automated, or are on the verge of, such as reading X-ray plates. We have organized a couple here [26]. Another benefit could actually reducing be costs: one benefice of AI models is that intelligence starts to be cheaper, once the model learns, it can be easily shared as API, as an example. Different from human intelligence, machine intelligence can be shared easily, and costless.

Humans, as Daniel Kahneman and colleagues [24] highlight, believe they understand how they decide, and get over-confident. It is not unknown about medical errors in diagnosis, and some areas of medicine may have a strong variation in the same diagnosis, coming from different professionals. [22] compared the diagnosis of medical doctors with machine learning using Optical coherence tomography (OCT): they have shown that this variation mentioned on humans compared to algorithms is real.

Machine learning, when make mistakes, it is easily traceable, and enhanced. Even though we do not understand precisely how those models learn, we do understand the big picture; in fact, [22] monitored where “the model was look at” on OCT for diagnostic, and it was correct. This is important to know when we are talking about models being used, and continuously enhanced based on their misdiagnosis. Once they learn, it does not happen again. Machine intelligence is easily repeatable, transferable and reliable once they are properly designed.

### 4.3. Mathematical models applied to medicine

We have essentially two big groups of models applied to medicine: white box and black models. The former focus on details, whereas the latter on the dataset.

Black box models are closer to how humans think, and decide. A goalkeeper can predict where the ball will most likely be, based on several cues/inputs, but he/she cannot explain it in details. You do not need to know all the physics from the ball to defend your goal. Machine learning is a black box model, and that is why they are so interesting to medicine. When we predict diabetes from a couple of features, it does not mean we understand the dynamics of diabetes, work done by white box models [16]. One can build a model for detecting a medical condition, from image or from physiological measurement without actually having a medical degree, or even, in-depth knowledge in medicine. This is something impressive about those models.

Most of the models in medicine, the ones called white box models, are complex, but not complex enough for reliable applications. Machine learning models, even though may get it right, are not explainable, they are just numbers that change during the learning process. Transfer learning is a set of numbers, which is used on another problem: knowledge from a set of images become a set of values on parameters called learning weights. They are just matrix multiplications and tensors manipulations. Different from humans when properly trained can explain their thinking in equations and theories, machine learning just do it right, but does not provide the rules explicitly.

Mathematical models have been applied to medicine for a long time, and eventually they become algorithms, which eventually may become software/packages. Those models can be classified in two big groups: white box models and black box models.

White box models are generally created for specific problems, and they concentrate on details. Their drawback is that they are specific, too much energy is expended on a model that can not be applied elsewhere, except for derivations from the problem solved. They are ideal for simulations and considering scenarios. On the other hand, black box models are generic, and not tied up to specific applications. The strong point is: if one area makes progress, it is automatically transferable to other areas. Those models do not consider applications, they are created based on general principles. TensorFlow,js, the technology we have used herein, was not designed for medicine, but it uses artificial neural networks as machine learning approach, which has been explored extensively to support medicine. This makes all the achievements elsewhere easily transferable, we have taken advantage of that observation herein.

### 4.4. Artificial intelligence as a prediction machine

All artificial intelligence nowadays is a prediction machine [27]: their goal is predicting what is next, based on a set of information called features. chatGPT is about prediction of what is the next word on a given text [28], we can predict snakes based on their details from an image [18], and we have done some predictions about medical conditions, such as diabetes. based on given features.

According to [27], those prediction machines can be compared to electricity and computer power: as they became cheaper and more accessible, new possibilities became real. Nowadays, those prediction machines are becoming cheaper, most of the time free, and easily used (e.g., pretrained models, APIs and public libraries). We have explored it herein, using a public library called TensorFlow.js, making the point that those prediction machines can be widely used, by a broad audience, no cost at all; and with all their capabilities and power to predict. We have already made the point on another article for image classification applied to biology [18], using another platform called Teachable Machine, which is built on top of TensorFlow.js. [22] applied to medical images those models, with success comparing to human experts.

### 4.5. On the importance of JavaScript as a computer language in machine learning

One group of researchers that have been changing medicine a lot in the last decades are computer scientists; and those models are a direct example. Computers are everywhere in medicine, and with the rise of artificial intelligence and computer power becoming cheaper and more accessible, it will accelerate this change (e.g., chatGPT in medicine [29]).

The point is: we need to make those apps available, as user-friendly as possible, as easily accessible as possible [30]. An informal survey showed that model parametrization (i.e., getting a model to execute a specific task) is the biggest issues amongst life scientists working with software [31]. Thus, it is something we must consider heavily when designing our apps: we should strongly think of the final end of the cycle, the user! We have explored with success using those model as a chatbot, where the artificial intelligence handles all [*to be published*].

Java was the very first computer language built to serve the browser, and it had its moment; the name JavaScript was given to call attention from the Java users. So far, all computer languages (e.g., C++, C, Fortran) were designed for desktop applications, to be used locally, on the user’s computers; they needed installation and constant maintenance; it was common to make software available as a command prompt. They were what is now called synchronous programming: the order of your lines of codes will dictate how it is executed, the user had no saying on this paradigm applied to coding. So, it came the idea of asynchronous programming: the user will interact with the program, and dictate how the code will evolve; to be fair, Java also had buttons and more, called listeners. The code will wait for user’s interaction, which triggers a set of lines of actions, which by themselves can be asynchronous.

It gave rise to multithreading: the ability of a code to handle more than one user at a time without requiring multiple copies of the program running on the computer, without blocking the main thread. It allowed also multiple tasks running simultaneously, and the app will still respond to the user normally; the actions go to background, and once it is finished, they give back the results.

We had our own experience with Java for creating applications for biomedical cases [32]. Our experience showed a couple of setbacks: i) graphs (e.g., result plotting) and ii) user interfaces (e.g., UI and UX) are hard to design; not to mentions other setbacks such as string manipulation.

We believe that similar setbacks will appear in any desktop computer languages, that includes Python. It is true that we can nowadays build a whole app using different languages, and we have done that [6]. The solution found by Python programmers, as an example, is using frontend frameworks/libraries (e.g., Django and Angular). The main issue with this workaround is having to use several languages and keeping different servers; for sure, you will need a bigger team, which increases costs, and difficulties to execute the project. Heroku, as an example, provides a possibility to easily build a pipeline based on different apps; thus, they work as one big program from the outside. Notwithstanding, still having to handle several apps in several languages.

In our case when dealing with these issues of having codes in different languages serving the same problem, we have used Galaxy [6]. It does not matter which choice you make, one may be better than other, they still require handling servers, and different codes in different language. The easily seen disadvantage of that is a bigger team; we all know that most research groups operate under zero or no research funding at all, not to say that actually making money from those scientific apps is a challenge. In our case [6], it made sense to choose Galaxy since the codes were already built by previous researchers, and rebuilding would be hard. The case we present herein is interesting mainly if you are starting, or have reasons to migrate to JavaScript. If you are starting a new project that requires machine learning, we recommend starting with JavaScript, and you will be able to work in a startup paradigm [33]: test it fast, as fast as possible. Not sure it is a good strategy to rebuild a complex app from scratch, unless you have reasons to believe it will improve.

Recently, thanks to several libraries, JavaScript became a viable option for designing machine learning. Nowadays, the most well-accepted and showing promising results is TensorFlow.js [1, 2, 3]. Thus, incorporate all the advantages of working with JavaScript and machine learning in one web application.

For our case, the core advantages are: i) the data will never leave the browser, ideal for sensitive data, most likely the case for medical information; ii) the calculations are done on the browser, no need to buy expensive server for scientific computations, ideal for startups and similar experimental endeavours. We believe that startups may support on decreasing the ever-growing costs of medical assistance [4]. They have already done that with text processing using artificial intelligence (e.g., chatGPT).

### 4.6. Diabetes and Smoking history

For making this statistical analysis, we have used the openAI Assistant [12].

The full dataset contains information about diabetic patients and includes the following columns:

1. gender: Gender of the patient (e.g., Male, Female)
2. age: Age of the patient
3. hypertension: Whether the patient has hypertension (1 for yes, 0 for no)
4. heart_disease: Whether the patient has heart disease (1 for yes, 0 for no)
5. ***smoking_history***: Smoking history of the patient (e.g., never, current, No Info)
6. bmi: Body Mass Index (BMI) of the patient
7. HbA1c_level: HbA1c level of the patient
8. blood_glucose_level: Blood glucose level of the patient
9. diabetes: Whether the patient has diabetes (1 for yes, 0 for no)

From the analysis and visualization, we can see that there are different proportions of diabetic patients within each category of smoking history. The proportions are as follows:

- No Info (Smoking History Not Available): 4.06%
- Current Smoker: 10.21%
- Ever Smoked: 11.79%
- Former Smoker: 17.00%
- Never Smoked: 9.53%
- Not Current Smoker: 10.70%

#### Obs

the proportion is the percentage of diabetic per category. For example, within the “never smoked” category, 9.53% of individuals have been diagnosed with diabetes.

Based on these proportions, we can see that former smokers have the highest proportion of diabetic patients, followed by those who “ever” smoked. To test the hypothesis that smoking may increase the chances of diabetes, we can perform a statistical test to compare the proportions of diabetic patients between different smoking history categories. We can use a chi-squared test to determine if there is a significant association between smoking history and diabetes. Let’s perform the chi-squared test to evaluate this hypothesis.

The p-value obtained from the chi-squared test is extremely small (approximately 0), indicating that *there is a statistically significant association between smoking history and diabetes*. This result suggests that smoking may indeed increase the chances of diabetes.

##### 4.6.1. Details on the hypothesis test

In the hypothesis test performed, the null hypothesis tested against is that there is no association between smoking history and the likelihood of having diabetes. In other words, the null hypothesis assumes that the proportions of diabetic patients are independent of the smoking history categories.

The rejection of the null hypothesis (as indicated by the extremely small p-value) suggests that there is a significant association between smoking history and diabetes. Therefore, we have evidence to suggest that smoking history may have an impact on the likelihood of having diabetes.

The chi-squared test was chosen because it is a statistical test commonly used to determine whether there is an association between categorical variables. In this case, we were interested in understanding if there is a significant association between smoking history (a categorical variable) and the likelihood of having diabetes (another categorical variable).

The chi-squared test is specifically designed to assess independence between two categorical variables by comparing the observed frequencies in a contingency table to the frequencies that would be expected if the variables were independent. If the calculated chi-squared statistic from the test is sufficiently large and the associated p-value is small, it suggests that there is a significant association between the two variables.

Given that we were testing the hypothesis that smoking may increase the chances of diabetes, the chi-squared test was deemed appropriate for this scenario to determine if there is a statistically significant association between smoking history and diabetes status in the dataset.

## Notes

### Competing Interest Statement

The authors have declared no competing interest.

### Funding Statement

This study did not receive any funding. It was produced independent by Jorge Guerra Pires at the project IdeaCoding Lab

### Author Declarations

The study used (or will use) ONLY openly available human data that were originally located at: Kaggle

### Summary of Updates

This is the 2rd round of the review process. Several changes were made upon suggestion from the reviewers, new simulations, I have written the text, and add new resources.

## References

[1] Rivera, J. D. D. S. Practical Tensor-Flow.js: Deep Learning in Web App Development.https://www.amazon.com/Practical-TensorFlow-js-Deep-Learning-Development/dp/1484262727: Apress, 2020. 328 p.

[2] Cai, S. B. S.; Nielsen, E. D.; Chollet, F. Deep Learning with JavaScript: Neural net-works in TensorFlow.js. [S.l.]: Manning, 2020. 560 p. Available at https://www.amazon.com/Deep-Learning-JavaScript-networks-TensorFlow-js/dp/1617296171⟩.

[3] Laborde, G. Learning Tensorflow.Js: Powerful Machine Learning in JavaScript. https://www.amazon.com.br/Learning-Tensorflow-Js-Powerful-Machine-JavaScript/dp/1492090794: O’Reilly Media, 2021. 338 p.

[4] Pires, J. G. Alguns insights em startups um novo paradigma para a tríplice aliança ciência, tecnologia e inovação: a novel paradigm for understanding the triple alliance of science, technology and innovation. Revista Gestão amp; Saúde, v. 11, n. 1, p. 38–54, fev. 2020. Disponível em: ⟨https://periodicos.unb.br/index.php/rgs/article/view/28626⟩.

[5] Fain, Y.; Moiseev, A. Angular Development with TypeScript. https://www.amazon.com/Angular-Development-Typescript-Yakov-Fain/dp/1617295345: Manning, 2018. 560 p.

[6] Pires, J. G. et al. Galaxy and mean stack to create a user-friendly workflow for the rational optimization of cancer chemotherapy. 2021. Disponível em: ⟨doi:10.3389/fgene.2021.624259⟩.

[7] Harman, L. B.; Flite, C. A.; Bond, K. Electronic health records: Privacy, confidentiality, and security. AMA Journal of Ethics, v. 14, n. 9, p. 712–719, 2012. Disponível em: ⟨https://journalofethics.ama-assn.org/article/electronic-health-records-privacy-confidentiality-and-security/2012-09⟩.

[8] Strauss, B. How to Safely Collect and Store Patient Data. 2020. Accessed: 2023-12-18. Disponível em: ⟨https://www.egnyte.com/blog/post/how-to-safely-collect-and-store-patient-data⟩.

[9] Haykin, S. Neural Networks and Learning Machines. 3rd. ed. [S.l.]: Pearson, 2008.

[10] FULL interview: “Godfather of artificial intelligence” talks impact and potential of AI. CBS Mornings, 2023. Accessed on 19 Dec 2023. Disponível em: ⟨https://www.youtube.com/watch?v=qpoRO378qRY⟩.

[11] Kistler, W. M.; Gerstner, W. Spiking Neuron Models: Single Neurons, Populations, Plasticity. [S.l.]: Cambridge University Press, 2002.

[12] Pires, J. G. Data Science using openAI: testing their new capabilities focused on data science. [S.l.]: Qeios, 2023. Preprint.

[13] Chen, W. et al. Nonlinear modeling using support vector machine for heart rate response to exercise. In:. Computational Intelligence and Its Applications. [s.n.]. p. 255–270. Disponível em: ⟨https://www.worldscientific.com/doi/abs/10.1142/97818481669290010⟩.

[14] Ling, S.; San, P.; Nguyen, H. Hypoglycemia detection for insulin-dependent diabetes mellitus: Evolved fuzzy inference system approach. In:. Computational Intelligence and Its Applications. [s.n.]. p. 61–85. Disponível em: ⟨https://www.worldscientific.com/doi/abs/10.1142/97818481669290004⟩.

[15] Alty, S.; Lam, H.; Prada, J. On the applications of heart disease risk classification and hand-written character recognition using support vector machines. In:. Computational Intelligence and Its Applications. [s.n.]. p. 213–253. Disponível em: ⟨https://www.worldscientific.com/doi/abs/10.1142/97818481669290009⟩.

[16] Palumbo, P. et al. Mathematical modeling of the glucose–insulin system: A review. Mathematical biosciences, v. 244, 05 2013.

[17] Campagna, D. et al. Smoking and diabetes: dangerous liaisons and confusing relationships. Diabetology Metabolic Syndrome, v. 11, n. 1, p. 85, 2019.

[18] Pires, J. G. Snakeface: a transfer learning based app for snake classification. bioRxiv, Cold Spring Harbor Laboratory, 2023. Disponível em: ⟨https://www.biorxiv.org/content/early/2023/06/14/2023.06.13.544741⟩.

[19] Lusignan, S. de et al. Miscoding, misclassification and misdiagnosis of diabetes in primary care. Diabetic Medicine, v. 29, n. 2, p. 181–189, 2012. Disponível em: ⟨https://onlinelibrary.wiley.com/doi/abs/10.1111/j.1464-5491.2011.03419.x⟩.

[20] Sehrawat, T. et al. Utility and limitations of glycated hemoglobin (hba1c) in patients with liver cirrhosis as compared with oral glucose tolerance test for diagnosis of diabetes. Mathematical biosciences, v. 9, p. 243–251, 2018.

[21] Meehl, P. E. Clinical Versus Statistical Prediction: A Theoretical Analysis and a Review of the Evidence. [S.l.]: Echo Point Books Media, 2013.

[22] Kermany, D. S. et al. Identifying medical diagnoses and treatable diseases by image-based deep learning. Cell, v. 172, n. 5, p. 1122–1131.e9, 2018. Disponível em: ⟨10.1016/j.cell.2018.02.010⟩.

[23] Cercato, C.; Fonseca, F. Cardiovascular risk and obesity. Diabetology & Metabolic Syndrome, Springer, v. 11, n. 1, p. 74, 2019.

[24] Sunstein, C. R.; Kahneman, D.; Sibony, O. Noise: A Flaw in Human Judgment. [S.l.]: Little, Brown Spark, 2021).

[25] Holmes, S.; Harber, C. Getting MEAN with Mongo, Express, Angular, and Node. Shelter Island, NY: Manning Publications, 2019.

[26] Pires, J. G. Relatório Final de pós-doutorado Programa Nacional de Pós-doutorado PNPD/CAPES. [S.l.], 2018. Disponível em: ⟨https://www.researchgate.net/publication/329815289 Relatorio Final de pos-doutorado Programa Nacional de Pos-doutorado PNPDCAPES⟩.

[27] Gans, J.; Goldfarb, A. Prediction Machines: The Simple Economics of Artificial Intelligence. [S.l.]: Harvard Business Review Press, 2018.

[28] Wolfram, S. What Is ChatGPT Doing … and Why Does It Work? https://www.amazon.com/What-ChatGPT-Doing-Does-Work/dp/1579550819: Wolfram Research, 2023. 112 p.

[29] Pires, J. G. Could chatGPT play a medical doctor? 2023. Published in Computational Thinking: How computers think, decide and learn. Disponível em: ⟨https://medium.com/computational-thinking-how-computers-think-decide/could-chatgpt-play-a-medical-doctor-8dfcd8c95538⟩.

[30] Pires, J. G. Innovating with biomathematics: the challenge of building user-friendly interfaces for computational biology. Academia Letters, 2022. Disponível em: ⟨10.20935/AL5792⟩.

[31] Pires, J. G. An informal survey presents the gap between computer and medical doctors and biologists. 2023. Published in Theoretical and Mathematical Biology (blog in Medium publication). Disponível em: https://bit.ly/3XfhCKH⟩.

[32] Pires, J. G. Biomechanics, Computational Intelligence, and Systems Biology with application on Vitreous Dynamics Using Java: an incipient discussion. 2014. Preprint in Academia Edu. Disponível em: ⟨https://www.academia.edu/8341040/Title Biomechanics Computational Intelligence and Systems Biology with application on Vitreous Dynamics Using Java an incipient discussion⟩.

[33] Blank, S.; Dorf, B. The Startup Owner’s Manual: The Step-By-Step Guide for Building a Great Company. Illustrated edition. [S.l.]: Wiley, 2020. 608 p.

